# Analysis of a Pilot Study Delivering Cancer Survivorship Education to Community Healthcare Professionals Utilizing the Project ECHO model

**DOI:** 10.1101/2023.09.25.23296103

**Authors:** Ashley Pariser, Kevin Johns, Dena Champion, Andrea Roberts, Susan Fugett, Erin Holley, Candice Schreiber, Carolyn J. Presley, Jalyn Todd, Andrew Honeychuck, Katherine Hunt, Yurong Lu, Bhuvaneswari Ramaswamy, Seuli Bose Brill

**Affiliations:** The Ohio State University Comprehensive Cancer Center, Columbus, OH; The Ohio State University Wexner Medical Center, Columbus, OH; The Ohio State University College of Medicine, Columbus, OH; Bon Secours St. Francis Cancer Center, Greenville, SC; Mercy Health St. Rita’s Cancer Center, Lima, OH; The Ohio State University Department of Statistics, Columbus, OH

**Keywords:** Cancer Survivorship, Project ECHO, Community Providers, Telehealth, Continuing Education

## Abstract

**Purpose:** This pilot study evaluated a 12-week Cancer Survivorship curriculum delivered utilizing the Project Echo® model on provider self-efficacy (SE), knowledge (KN), and professional improvement (PI).

**Methods:** Providers affiliated with the Mercy Health System were enrolled in two cohorts. Six one-hour sessions were developed from a needs assessment and delivered over 12 weeks. Participants completed pre and post session surveys evaluating 3 domains: SE, KN and PI. The average score for survey items overall and within each domain was compared pre- and postsurvey results.

**Results:** Twenty-six participants completed the baseline survey and 22 completed the post-study survey. For cohort 1, the overall score (0.94 (0.45,1.42) (P=0.0023), SE (1.1 (0.5,1.7) p = 0.003), and KN domain (1.03 (0.45,1.62) p= 0.0036) scores significantly increased. For cohort 2, the overall score (0.617 (0.042,1.193) p=0.0378), the SE (0.728(0.048,1.407), p = 0.0379), and KN domains (0.665 (0.041,1.289), p= 0.0387) increased significantly. The PI did not change for either cohort.

**Conclusions:** This Cancer Survivorship ECHO pilot resulted in a statistically significant increase in provider self-efficacy and knowledge. All 22 participants rated the Cancer Survivorship ECHO pilot experience as a positive (greater than neutral) on their training.

**Implications for Cancer Survivors:** The Cancer Survivorship ECHO model may serve as a scalable strategy for building cancer survivorship care capacity in community-based oncology practices through equipping multidisciplinary teams to meet the needs of cancer survivors within their community. Further research is needed to assess implementation of this model into novel settings and evaluate its impact on patient outcomes.

## Introduction

In 2023, there will be approximately 2 million new cases of cancer with 74,140 cases in the state of Ohio. As 1 in 2 men and 1 in 3 women are expected to be diagnosed with invasive cancer within their lifetime and 5-year relative survival for all cancers continues to improve, the need for access to guideline-based survivorship cancer care also continues to rise.[1] There is expected to be a significant increase in patients living 5 years and beyond their diagnosis, survivors older than age 65, as well as, a rise in patients living with stage IV disease.[2] Despite known short and long-term needs among a heterogeneous cancer survivorship population, access to guideline-based survivorship care remains limited.[3]

Unfortunately, in the United States, there are multiple barriers to expanding cancer care to meet the demands of patients receiving active and survivorship treatment alike including cost, work force shortages, and increasing complexity of both care and care coordination.[4] There are known cancer outcome disparities for patients living in rural areas compared to patients living in urban areas. Access to healthcare infrastructure, oncologic support services, and education are just 3 known inequities facing patients living with cancer in rural areas.[5]

Although multiple models for survivorship care delivery exist and the ideal survivorship model has yet to be elucidated, specialist-led post-treatment models in academic settings remain predominant.[3, 6] [7, 8] With only 21% of all oncology practices based in the academic setting and studies showing that upwards of 24% of all Medicare beneficiaries travel longer than an hour for their oncologic care, access to survivorship care in the United States remains unequal.[9, 10]

A known barrier to implementation of other survivorship models (i.e. led by primary care physicians, shared care) is a perceived lack of cancer-specific knowledge and skills.[11] Enhancing continuing education for non-academic oncology providers and non-oncology providers has been listed by multiple studies as well as professional societies as means to enhance implementation of survivorship care.[6, 11, 12] Project ECHO® is a hub and spoke model that utilizes a web-based platform (Zoom) for delivery of content through short didactics and case-based discussions. Project ECHO® was initially developed to enhance access to hepatitis C treatment for patients living in New Mexico.[13] Since its inception, it has been adopted and modified to enhance training of healthcare professionals in a diverse array of community settings. Currently highlighted on the Project ECHO Cancer website, there are 108 Hubs in 27 countries utilizing the Project ECHO model to engage community providers in topics related to cancer prevention, cancer screening or cancer treatment.[14]

Our pilot study highlights a collaboration between an academic comprehensive cancer center in Central Ohio and a multistate network - Bon Secours Mercy Health. This pilot study is the first to highlight the potential of an academic-community collaboration to enhance cancer survivorship education utilizing the Project ECHO model to meet site-specific educational needs of community oncology providers. Furthermore, it highlights the potential of such partnerships to continue to enhance access to educational opportunities attuned to provider, site, and community-specific needs.

## Methods

This cohort pilot study evaluated a 12-week Cancer Survivorship curriculum delivered utilizing the Project Echo® model on provider self-efficacy (SE), knowledge (KN), and professional improvement (PI).

This pilot was created in partnership with research leadership at Bon Secours-Mercy Health. This facilitated site identification optimizing recruitment of interested sites with ability to provide protected time to participants. Community oncology providers were recruited from two community hospitals within a multistate network -Bon Secours Mercy Health. As survivorship care requires a team approach, all providers involved in direct patient care were invited to participate.[3] This is in alignment with our previous work as well as other Project ECHO sites that enrolled participants with diverse professional backgrounds.[15] Enrolled participants included the following occupations: physicians, advanced practice providers, nurses, social workers, patient navigators, clinical research coordinator, pharmacist, oncology quality supervisor. Informed consent was obtained from all individual participants in this study.

A previously developed needs assessment based on ASCO guidelines, NCCN guidelines and aligned with the four established tenets of survivorship care was utilized.[16] The needs assessment was delivered securely via RedCap to interested, eligible participants at each cohort site. The needs assessment invited participants to indicate level of interest on a 5-point Likert scale ranging from very uninterested to very interested in 18 cancer survivorship topics. The following are examples of included topics: management of cancer related fatigue, management of chemotherapy induced neuropathy, and management of anxiety and depression in cancer survivors. The six topics of highest interest for each cohort were selected for the Cancer Survivorship ECHO curriculum.

For each cohort, six 1-hour sessions were delivered via Zoom over a 12-week period. The format of each session was aligned with the Project ECHO model and consisted of a didactic followed by case discussion. Didactics were presented and case discussions were moderated by a multidisciplinary panel. The multidisciplinary panel had representation from the following specialties: medical oncology, psychiatry, social work, nutrition, and physical therapy. As appropriate, additional specialists were recruited for individual sessions to enhance expertise on the multidisciplinary panel. All experts were involved in didactic content development.

All consented participants were invited to complete a pre and post survey. We developed the survey from surveys previously validated by the Project ECHO Institute. These surveys were amended to address provider’s knowledge and comfort in key survivorship competencies. Differences in participants pre- and post-intervention surveys determined change in provider comfort and knowledge with core cancer survivorship competencies. Areas assessed in the survey include professional satisfaction, patient access to survivorship care, provider ability to identify and counsel patients on survivorship care as well as knowledge and ability to address the four core tenets of survivorship care with patients. This survey evaluated for self-efficacy (SE) in cancer survivorship care, knowledge (KN) of cancer survivorship care, and professional improvement (PI). Each question consisted of a 5-point Likert scale with 1 reflecting lowest level of agreement, confidence, or knowledge and 5 reflecting highest level of agreement, confidence, or knowledge. To optimize survey collection, RedCap was utilized for secure delivery and collection and a gift card was offered to all participants for completion of both the pre- and post-surveys. In addition to the pre/post survey, an option for formative feedback was provided on each post-session continuing medical education (CME) survey.

### Statistical Analysis

Likert scale characteristics were summarized using frequencies (proportions), and character ordinal categories for each survey question were recoded to numerical values. All character ordinal categories were recorded to numerical values. Linear mixed models were used for the average score comparisons between pre and post survey for self-efficacy, knowledge domains, professional improvement and for overall comparison as well. A p value equal to or less than 0.05 was considered statistically significant. The statistical analysis for this study was performed in Statistical Analysis System (version 9.4, SAS Inst Inc, NC, USA) with proc mixed procedure. Comparison figures were accomplished in RStudio (R version 4.2.1 (2022-06-23 ucrt).

## Results

A total of 28 participants enrolled with 14 consented participants in each cohort. The first cohort included providers at a stand-alone, community oncology practice in Greenville, South Carolina. The following occupations were represented in cohort 1: physician (1), advanced practice provider (3), nurse (4), social worker (2), dietitian (1), clinical research coordinator (1), pharmacist (1), oncology quality supervisor (1). The second cohort included providers at a community oncology practice affiliated with a community hospital in Lima, Ohio. Cohort 2 had the following breakdown by occupation: advanced practice provider (3), nurse (8), dietitian (1), patient navigators (2). With exception of one session, all sessions had ≥85% attendance.

Topics of highest interest on the needs assessment were prioritized and selected for curriculum development and delivery. Both cohorts prioritized the following three topics: Adjusting to and Preparing Patients for Survivorship (Survivorship 101), Cancer Related Fatigue, and Anxiety and Depression. Additional topics selected by cohort 1 included Chemo Brain, Neuropathy, and Sexual Health, while cohort 2 selected Risk of Recurrence, Nutrition Myths, and Bone Health.

All participants were invited to provide formative feedback on continuing medical education (CME) surveys provided at the conclusion of each session. The multidisciplinary panel met periodically throughout the study to reflect and assess program development and implementation. During cohort 1, there was minimal active participation from enrolled participants. The bulk of discussion and all cases were presented by members of the multidisciplinary panel. This resulted in implementation of more active learning strategies during cohort 2. For example, the sign-in list was utilized to encourage and invite all participants of the session to provide feedback on didactics, case discussions and embedded group think-share opportunities. In contrast to cohort 1, all but one case discussion was provided and presented by cohort 2 participants.

Data from both pre- and post-surveys was available for 22 of 28 enrolled participants (78.5%). For cohort 1, there were 13 participants who completed the survey for this pilot study. One did not complete the presurvey and 3 did not complete the postsurvey. The average scores for overall score and each domain (SE, KN, PI) are summarized in **Table 1**. The average overall score increased significantly 0.94 (0.45, 1.42) (P=0.002). Additionally, average score for SE and KN domains increased significantly: SE (1.10 (0.50,1.70) p=0.003 and KN (1.03) (0.45, 1.62) p=0.004. PI did not change likely reflective of a ceiling effect given pre-survey score of 4.542 ± 0.582.

**Table 1.** Survey Results for Cohort 1: Professional Improvement (PI), Self-Efficacy (SE), Knowledge (KN), and Overall Score.

|  |  | Presurvey |  |  | Postsurvey |  |  | Changes* |  |
| --- | --- | --- | --- | --- | --- | --- | --- | --- | --- |
|  |  | N | Mean | STD | N | Mean | STD | mean (95% CI) | P values |
| PI | (2Qs) | 12 | 4.542 | 0.582 | 10 | 4.450 | 0.599 | -0.09 (-0.67,0.49) | 0.726 |
| SE | (8Qs) | 12 | 2.917 | 0.679 | 10 | 4.013 | 0.508 | 1.10 (0.50,1.70) | 0.003 |
| KN | (8Qs) | 12 | 2.531 | 0.681 | 10 | 3.563 | 0.465 | 1.03 (0.45,1.62) | 0.004 |
| Overall | (18Qs) | 12 | 2.926 | 0.563 | 10 | 3.861 | 0.399 | 0.94 (0.45,1.42) | 0.002 |

\* based on linear mixed model for repeated data, SAS software (version 9.4, SAS Inst Inc, NC, USA) proc Mixed procedure

For Cohort 2, 14 participants completed a survey for this study. Two participants did not complete the post-survey. The average scores for each domain (SE, KN, PI) are summarized in **Table 2**. The overall average score, SE, and KN all increased significantly from presurvey to postsurvey. Change in mean score were 0.62 (0.04, 1.19) (p=0.038), 0.73(0.05, 1.41) (p=0.038), and 0.67 (0.04, 1.29) (p=0.039) respectively. Average PI score did not change from pre- to postsurvey, 0.00 (-0.79, 0.79) (p≥0.999).

**Table 2.** Survey Results for Cohort 2: Professional Improvement (PI), Self-Efficacy (SE), Knowledge (KN), and Overall Score.

|  | Presurvey | Postsurvey | Changes* |
| --- | --- | --- | --- |

|  |  | N | Mean | STD | N | Mean | STD | mean (95% CI) | P values |
| --- | --- | --- | --- | --- | --- | --- | --- | --- | --- |
| PI | (2Qs) | 14 | 4.000 | 0.941 | 12 | 4.000 | 0.879 | 0.00 (-0.79,0.79) | > 0.999 |
| SE | (8Qs) | 14 | 3.366 | 0.923 | 12 | 4.094 | 0.579 | 0.73(0.05,1.41) | 0.038 |
| KN | (8Qs) | 14 | 2.991 | 0.791 | 12 | 3.656 | 0.626 | 0.67 (0.04,1.29) | 0.039 |
| Overall | (18Qs) | 14 | 3.272 | 0.735 | 12 | 3.889 | 0.571 | 0.62 (0.04,1.19) | 0.038 |

\* based on linear mixed model for repeated data, SAS software (version 9.4, SAS Inst Inc, NC, USA) proc Mixed procedure

The postsurvey included 4 questions that evaluated the impact on ECHO on participants’ professional isolation, satisfaction, and patient access to care (**Table 3**). For cohort 1, a total of 10 participants completed the post-survey. For cohort 2, a total of 12 participants completed the post-survey. All 22 participants agreed that there was a positive effect (average score of 3.4 for cohort 1 and 3.8 for cohort 2) of the Cancer Survivorship ECHO on their training.

**Table 3.** Summary for Postsurvey Only Questions.

| Postsurvey Questions Only | Cohort 1 |  |  |  |  | Cohort 2 |  |  |  |  |
| --- | --- | --- | --- | --- | --- | --- | --- | --- | --- | --- |
|  | N | Mean | Q1 | Median | Q3 | N | Mean | Q1 | Median | Q3 |
| ECHO has diminished my professional isolation | 10 | 3.7 | 3.0 | 4.0 | 4.0 | 12 | 3.8 | 3.5 | 4.0 | 4.0 |
| My participation in ECHO has enhanced my professional satisfaction. | 10 | 3.9 | 3.8 | 4.0 | 4.3 | 12 | 4.6 | 4.0 | 5.0 | 5.0 |
| My participation in ECHO is a benefit to my place of employment. | 10 | 4.1 | 3.8 | 4.0 | 5.0 | 12 | 4.8 | 4.5 | 5.0 | 5.0 |
| ECHO has expanded access to Cancer Survivorship care for patients in our community. | 10 | 3.4 | 3.0 | 3.0 | 4.0 | 12 | 3.8 | 3.0 | 4.0 | 4.0 |

Except for PI, all postsurvey scores were higher than presurvey scores for both cohort 1 and cohort 2. The changes in score for overall, SE, and KN were all statistically significant. Equality reference lines were calculated for both cohort 1 and 2 based on number of participants who completed both pre and post surveys.

## Discussion

The Project ECHO model utilizes a low-cost, technology-based hub and spoke model embedded in grounded and situated learning theory. It has the potential to rapidly disseminate knowledge and expertise to providers throughout the world.[17-20] Our pilot study highlights the successful implementation of a new Project ECHO hub in Central Ohio and builds upon growing body of literature demonstrating the feasibility of the Project ECHO model to enhance provider knowledge in guideline-based cancer survivorship practices.[15, 16, 21, 22] Our study was able to demonstrate successful implementation of Moore’s framework 1-4: Participation, Satisfaction, Learning and Competence as demonstrated by a statistically significant increase in overall, selfefficacy, and knowledge domains in our survey results. [19]

**Figure 1:**
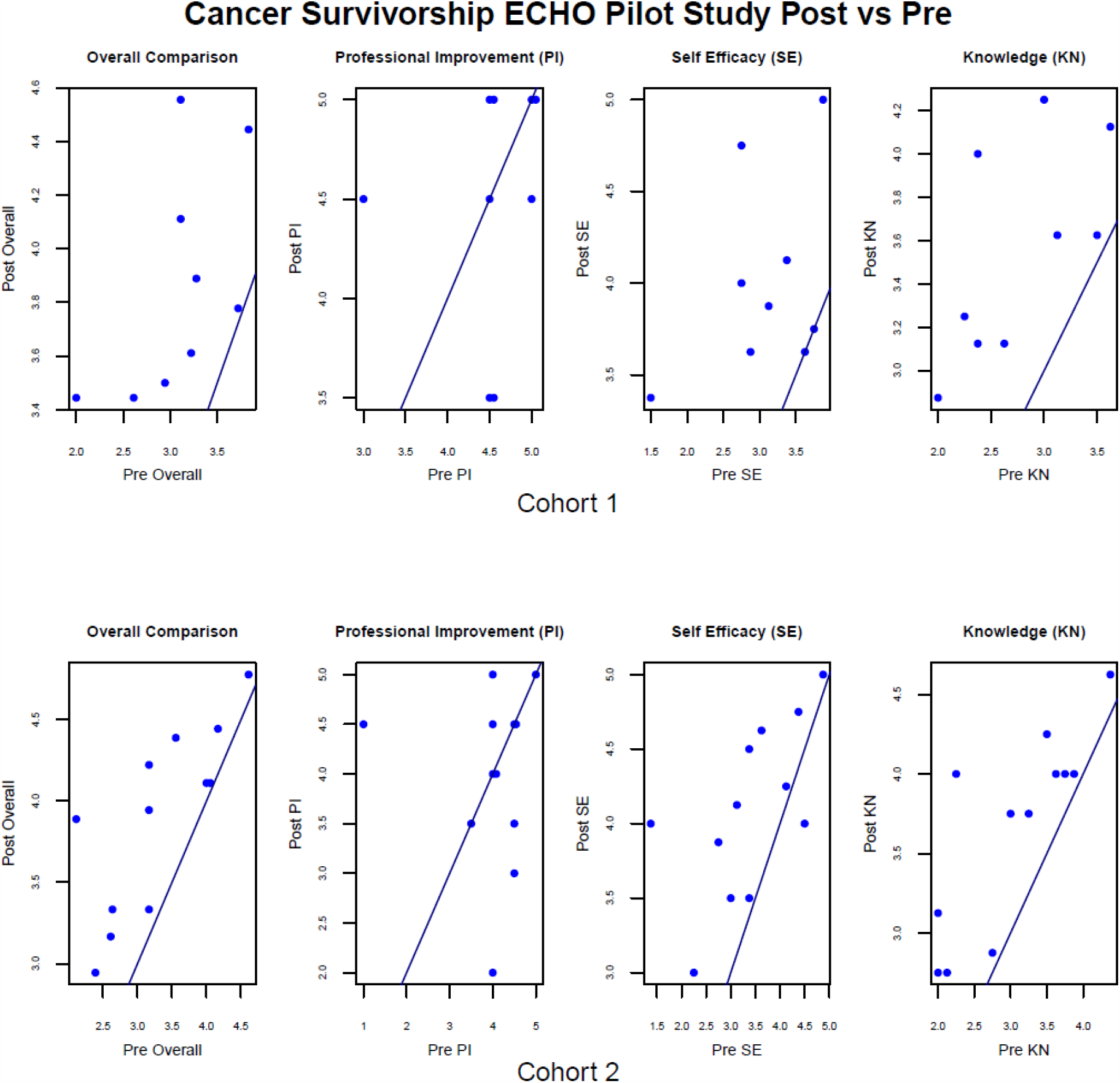
Participation in the Cancer Survivorship ECHO Pilot Study Yielded Improvement in Self-Efficacy, Knowledge, and Overall Comparison.

Furthermore, our pilot highlights the potential of academic-community partnerships to enhance recruitment and participation. A community co-design approach was utilized to align research and educational goals of between the hub and community sites. [23] Research leadership at Bon Secours Mercy Health and cohort site leaders were engaged and provided feedback from design through implementation of our study. This facilitated site identification optimizing recruitment of interested sites with ability to provide protected time to participants.

Our Cancer Survivorship ECHO builds upon previous work developing a novel, multidisciplinary Project ECHO hub.[16] Several modifications were made to address limitations of that previous study. These included enhanced protected time for participants, RedCap utilization for secure delivery and collection of surveys, continuing medical education (CME) credit for each attended session, and gift card offerings to all participants for completion of both the pre- and post-surveys. Our results demonstrate enhanced session attendance (greater than 85% for 11 of the 12 sessions) and availability of data from both pre- and post-surveys for enrolled participants (78.5%) because of these modifications.

Our post-survey findings suggest that Project ECHO may be a useful component for addressing clinician burnout and resourcing to address complex care needs. Post-surveys (Table 3) demonstrated mean scores greater than 3 for questions assessing this pilot’s impact on professional satisfaction and diminishing professional isolation. This highlights the potential of the Project ECHO model for enhancing support for providers and means of team building.[24] More research is needed to confirm replicability of this finding.

Our study had several limitations. First, as a pilot study of a novel Project ECHO hub in cancer survivorship, only two cohort sites were enrolled with a small, overall, sample size. Despite the small sample size, similarities of findings between cohort sites may indicate broader applicability of our study findings; however, future work is needed to confirm these findings in additional community sites. Second, cohort recruitment and delivery of Echo sessions were site based.

This site-specific approach had the potential advantages of team building and highlighting site and community specific resources. Furthermore, this approach allowed for buy-in from multiple key stakeholders including leadership. Our approach also resulted in excellent session attendance. To our knowledge, no study has evaluated efficacy of site-specific vs multi-site ECHO spoke model. With multiple demands on providers’ time, this approach may reduce barriers to participation and presents an opportunity for ongoing academic-community collaborations for enhancing access to survivorship care. Our team is conducting qualitative interviews to identify programmatic barriers and enhance survey components of program evaluation. This will allow for ongoing program evaluation and enhancement with future site recruitment and implementation.

Enhancing access to guideline-based survivorship care remains an important, unmet need. As cancer survivors are a heterogeneous and growing population with distinct short and long-term health needs, novel models and frameworks that allow for risk-based delivery of care that meet the individual needs of patients while also balancing known workforce shortages are needed.[25, 26] As cancer survivorship education has not been included in formalized training, our research seeks to enhance patient access to cancer survivorship care through recruitment, training, and support of diverse population of healthcare providers.

Our Cancer Survivorship ECHO pilot demonstrated the feasibility of utilizing a co-design approach to develop and implement a new Project ECHO program with improvement in provider self-efficacy, knowledge, and overall scores on pre/post surveys. While this pilot study recruited community oncology providers from two community sites, the Survivorship Cancer ECHO model has potential to be a central strategy for addressing survivorship workforce issues by increasing capacity within all community oncology settings. Additionally, this model may have broad applicability to other settings (i.e., primary care) to enhance capacity to meet the needs of all survivors including long-term and low-risk survivors. Future research in evaluating replicability in expanded sites as well as feasibility and patient outcomes will advance understanding of the utility of this model for enhancing survivorship care delivery. Applying our community-based codesign approach[23] will be critical to ensuring that future applications address local needs of stakeholders and will be central to the implementation strategy.

## Conclusions

Novel partnerships and team-based approach is feasible to rapidly disseminate knowledge to all providers engaged in direct patient care. Additional research accessing implementation of the Project ECHO platform into novel environments as well as ongoing assessments of impact of these programs on patient care outcomes is needed.

## Supporting information

Supplemental Needs Assessment

Supplemental Pre- and Post-Clinic Survey

## Data Availability

All data produced in the present study are available upon reasonable request to the authors.

## Notes

**Funding:** The project described was supported by a Path to K award (Grant # GR123938) from the Ohio State University Office of Health Sciences and the Center for Clinical & Translational Science. The content is solely the responsibility of the authors and does not necessarily represent the official views of the university, or the Center for Clinical and Translational Science.

### Competing Interest Statement

The authors have declared no competing interest.

### Clinical Trial

OSU21149

### Funding Statement

This study was supported by a Path to K award (Grant # GR123938) from The Ohio State University Office of Health Sciences and the Center for Clinical & Translational Science. The content is solely the responsibility of the authors and does not necessarily represent the official views of the university, or the Center for Clinical and Translational Science.

### Author Declarations

This study was performed in line with the principles of the Declaration of Helsinki. The Institutional Review Board of The Ohio State University approved this study.

## References

[1] R. L. Siegel, K. D. Miller, N. S. Wagle, and A. Jemal, “Cancer statistics, 2023,” CA: a cancer journal for clinicians, vol. 73, no. 1, pp. 17–48, 2023.

[2] N. C. Institute. “Statistics and Graphs.” National Cancer Institute Division of Cancer Control & Population Sciences. https://cancercontrol.cancer.gov/ocs/statistics (accessed 4.20.2023, 2023).

[3] C. M. Alfano, K. Oeffinger, T. Sanft, and B. Tortorella, “Engaging TEAM Medicine in Patient Care: Redefining Cancer Survivorship From Diagnosis,” American Society of Clinical Oncology Educational Book, vol. 42, pp. 921–931, 2022.

[4] K. Noyes, J. R. Monson, I. Rizvi, A. Savastano, J. S. Green, and N. Sevdalis, “Regional multiteam systems in cancer care delivery,” Journal of oncology practice, vol. 12, no. 11, pp. 1059–1066, 2016.

[5] S. Bhatia et al., “Rural–urban disparities in cancer outcomes: opportunities for future research,” JNCI: Journal of the National Cancer Institute, vol. 114, no. 7, pp. 940–952, 2022.

[6] R. J. Chan et al., “Effectiveness and implementation of models of cancer survivorship care: an overview of systematic reviews,” Journal of Cancer Survivorship, pp. 1–25, 2021.

[7] L. Nekhlyudov, “Integrating primary care in cancer survivorship programs: models of care for a growing patient population,” The oncologist, vol. 19, no. 6, pp. 579–582, 2014.

[8] M. T. Halpern, M. S. McCabe, and M. A. Burg, “The cancer survivorship journey: models of care, disparities, barriers, and future directions,” American Society of Clinical Oncology Educational Book, vol. 36, pp. 231–239, 2016.

[9] M. K. Kirkwood et al., “The State of Oncology Practice in America, 2018: Results of the ASCO Practice Census Survey,” Journal of Oncology Practice, vol. 14, no. 7, pp. e412–e420, 2018, doi: 10.1200/jop.18.00149.

[10] G. B. Rocque et al., “Impact of travel time on health care costs and resource use by phase of care for older patients with cancer,” Journal of Clinical Oncology, vol. 37, no. 22, p. 1935, 2019.

[11] K. Lisy, J. Kent, A. Piper, and M. Jefford, “Facilitators and barriers to shared primary and specialist cancer care: a systematic review,” Supportive Care in Cancer, vol. 29, pp. 85–96, 2021.

[12] I. Vaz-Luis et al., “ESMO Expert Consensus Statements on Cancer Survivorship: Promoting high-quality survivorship care and research in Europe,” Annals of Oncology, vol. 33, no. 11, pp. 1119–1133, 2022.

[13] S. Arora et al., “Outcomes of hepatitis C treatment by primary care providers,” The New England journal of medicine, vol. 364, no. 23, 2011.

[14] U. H. S. P. ECHO. “Cancer ECHO Revolutionizing Cancer Care Delivery.” https://hsc.unm.edu/echo/partner-portal/echos-initiatives/bmsf-cancer/ (accessed 4.20.2023, 2023).

[15] M. A. Etling et al., “The continuing evolution of a cancer prevention, screening, and survivorship ECHO: A second year of implementation,” Cancer Medicine, 2022.

[16] A. C. Pariser, J. Brita, M. Harrigan, S. Capozza, A. Khairallah, and T. B. Sanft, “Delivery of cancer survivorship education to community healthcare professionals,” Journal of Cancer Education, pp. 1–7, 2022.

[17] M. L. Varon et al., “Project ECHO cancer initiative: a tool to improve care and increase capacity along the continuum of cancer care,” Journal of Cancer Education, vol. 36, pp. 25–38, 2021.

[18] K. Guldberg, “Adult learners and professional development: peer-to-peer learning in a networked community,” International Journal of Lifelong Education, vol. 27, no. 1, pp. 35–49, 2008.

[19] D. E. Moore Jr, J. S. Green, and H. A. Gallis, “Achieving desired results and improved outcomes: integrating planning and assessment throughout learning activities,” Journal of continuing education in the health professions, vol. 29, no. 1, pp. 1–15, 2009.

[20] J. Lave and E. Wenger, Situated learning: Legitimate peripheral participation. Cambridge university press, 1991.

[21] J. Agley, J. Delong, A. Janota, A. Carson, J. Roberts, and G. Maupome, “Reflections on project ECHO: qualitative findings from five different ECHO programs,” Medical Education Online, vol. 26, no. 1, p. 1936435, 2021.

[22] C. Zhou, A. Crawford, E. Serhal, P. Kurdyak, and S. Sockalingam, “The impact of project ECHO on participant and patient outcomes: a systematic review,” Academic Medicine, vol. 91, no. 10, pp. 1439–1461, 2016.

[23] T. Zamenopoulos and K. Alexiou, Co-design as collaborative research. Bristol University/AHRC Connected Communities Programme, 2018.

[24] C. A. LeNoble, R. Pegram, M. L. Shuffler, T. Fuqua, and D. W. Wiper III, “To address burnout in oncology, we must look to teams: reflections on an organizational science approach,” JCO Oncology Practice, vol. 16, no. 4, pp. e377–e383, 2020.

[25] L. Nekhlyudov, M. A. Mollica, P. B. Jacobsen, D. K. Mayer, L. N. Shulman, and A. M. Geiger, “Developing a quality of cancer survivorship care framework: implications for clinical care, research, and policy,” JNCI: Journal of the National Cancer Institute, vol. 111, no. 11, pp. 1120–1130, 2019.

[26] C. M. Alfano et al., “Equitably improving outcomes for cancer survivors and supporting caregivers: a blueprint for care delivery, research, education, and policy,” CA: a cancer journal for clinicians, vol. 69, no. 1, pp. 35–49, 2019.

