## Supplemental Needs Assessment for "Analysis of a Pilot Study Delivering Cancer Survivorship Education to Community Healthcare Professionals Utilizing the Project ECHO model"

**OSU Survivorship Program ECHO- Clinic Needs Assessment Survey**

**For each of the following questions, rate your interest in learning about the following cancer survivorship education topics using the rating scale.**

|  | **Very Uninterested** | **Uninterested** | **Neutral** | **Interested** | **Very Interested** | **Prefer not to answer** |
| --- | --- | --- | --- | --- | --- | --- |
| 1. “Am I cancer free?”  How to communicate risks of recurrence. |  |  |  |  |  |  |
| 2. “What do I do now?”  How to discuss adjusting to new normal and survivorship. |  |  |  |  |  |  |
| 3. “Can I pass this on?”  When to refer to cancer genetics. |  |  |  |  |  |  |
| 4. “I know I should be happy, but…”  How to discuss and manage anxiety and depression in a cancer survivors. |  |  |  |  |  |  |
| 5. “Does sugar feed my cancer?” How to address nutrition myths and evidence. |  |  |  |  |  |  |
| 6. “But it’s natural?”  How to guide patients on do’s and don’ts of supplements. |  |  |  |  |  |  |
| 7. “Why can’t I lose weight?” How to discuss exercise and weight management. |  |  |  |  |  |  |
| 8. “I would if I had time.”  How to tailor lifestyle management and exercise. |  |  |  |  |  |  |
| 9. “I just can’t remember.” How to discuss and manage effects of chemo brain and memory loss. |  |  |  |  |  |  |
| 10. “Neuropathy and the cancer survivor.” Management and treatment strategies for treatment related neuropathy. |  |  |  |  |  |  |
| 11. “I’m just so tired.”  How to manage and support patients with cancer related fatigue. |  |  |  |  |  |  |
| 12. “Heart health and the cancer survivor.”  How to avoid cardiotoxicity. |  |  |  |  |  |  |
| 13. “Sex after Cancer.”  How to screen, discuss and manage sexual dysfunction. |  |  |  |  |  |  |
| 14. “Bone Health and the Cancer Survivor.”  Management strategies for avoiding osteoporosis. |  |  |  |  |  |  |
| 15. “Complementary Medicine and its Role in Survivorship.” Strategies and data of complementary medicine services. |  |  |  |  |  |  |
| 16. “What if I want a baby?” How to discuss infertility and pregnancy options. |  |  |  |  |  |  |
| 17. “Survivorship for Older Survivors.”  Strategies and implementation of geriatric assessments to optimize care for older cancer survivors. |  |  |  |  |  |  |
| 18. “I’m struggling to find resources that apply to me.”  Tips and strategies to augment delivery of culturally competent care to a diverse and heterogeneous population of cancer survivors. |  |  |  |  |  |  |
