## Supplemental Pre- and Post-Clinic Survey for "Analysis of a Pilot Study Delivering Cancer Survivorship Education to Community Healthcare Professionals Utilizing the Project ECHO model"

**Survivorship ECHO Project Provider Survey**

(Hidden Date: MM-DD-YYYY)

**Please answer each of the following questions.**

**Professional Improvement:**

1. ECHO has diminished my professional isolation. [Post survey only]
   a. Strongly Disagree
   b. Disagree
   c. Neutral
   d. Agree
   e. Strongly Agree
   f. Prefer not to answer
2. My participation in ECHO has enhanced my professional satisfaction. [Post survey only]
   a. Strongly Disagree
   b. Disagree
   c. Neutral
   d. Agree
   e. Strongly Agree
   f. Prefer not to answer
3. My participation in ECHO is a benefit to my place of employment. [Post survey only]
   a. Strongly Disagree
   b. Disagree
   c. Neutral
   d. Agree
   e. Strongly Agree
   f. Prefer not to answer
4. ECHO has expanded access to Cancer Survivorship care for patients in our community. [Post survey only]
   a. Strongly Disagree
   b. Disagree
   c. Neutral
   d. Agree
   e. Strongly Agree
   f. Prefer not to answer
5. In general, access to Cancer Survivorship expertise and consultation is a major area of need for me. [Pre and Post Survey]
   a. Strongly Disagree
   b. Disagree
   c. Neutral
   d. Agree
   e. Strongly Agree
   f. Prefer not to answer
6. Access to Cancer Survivorship expertise and consultation is a major area of need for my place of employment. [Pre and Post Survey]
   a. Strongly Disagree
   b. Disagree
   c. Neutral
   d. Agree
   e. Strongly Agree
   f. Prefer not to answer

**Self-Efficacy in Cancer Survivorship Care:** [Pre and Post Survey]

**This set of questions asks about your confidence in your abilities.**

1. On a scale of 1 to 5, how confident are you in your ability to address Cancer Survivorship care utilizing a multidisciplinary approach?
   a. 1 – Not confident
   b. 2 – Less confident
   c. 3 – Somewhat confident
   d. 4 - Confident
   e. 5 – Very confident
   f. Prefer not to answer
2. On a scale of 1 to 5, how confident are you in your ability to identify patients who would benefit from Cancer Survivorship care?
   a. 1 – Not confident
   b. 2 – Less confident
   c. 3 – Somewhat confident
   d. 4 - Confident
   e. 5 – Very confident
   f. Prefer not to answer
3. On a scale of 1 to 5, how confident are you in your ability to educate fellow health care providers about Cancer Survivorship care?
   a. 1 – Not confident
   b. 2 – Less confident
   c. 3 – Somewhat confident
   d. 4 - Confident
   e. 5 – Very confident
   f. Prefer not to answer
4. On a scale of 1 to 5, how confident are you in your ability to educate patients and families about Cancer Survivorship care?
   a. 1 – Not confident
   b. 2 – Less confident
   c. 3 – Somewhat confident
   d. 4 - Confident
   e. 5 – Very confident
   f. Prefer not to answer
5. On a scale of 1 to 5, how confident are you in your ability to assess and manage concerns that Cancer Survivors face?
   a. 1 – Not confident
   b. 2 – Less confident
   c. 3 – Somewhat confident
   d. 4 - Confident
   e. 5 – Very confident
   f. Prefer not to answer
6. On a scale of 1 to 5, how confident are you in your ability to manage and treat the multiple concerns that face cancer survivors?
   a. 1 – Not confident
   b. 2 – Less confident
   c. 3 – Somewhat confident
   d. 4 - Confident
   e. 5 – Very confident
   f. Prefer not to answer
7. On a scale of 1 to 5, how confident are you in your ability to communicate with cancer survivors and their families?
   a. 1 – Not confident
   b. 2 – Less confident
   c. 3 – Somewhat confident
   d. 4 - Confident
   e. 5 – Very confident
   f. Prefer not to answer
8. On a scale of 1 to 5, how confident are you in your ability to discuss Cancer Survivorship with peers?
   a. 1 – Not confident
   b. 2 – Less confident
   c. 3 – Somewhat confident
   d. 4 - Confident
   e. 5 – Very confident
   f. Prefer not to answer

**Knowledge of Cancer Survivorship Care:** [Pre and Post Survey]

**The next questions ask about how informed you feel about common aspects that concern cancer survivors.**

1. On a scale of 1 to 5, how much knowledge do you have of Cancer Survivorship care?
   a. 1 - No Knowledge
   b. 2 - Slight Knowledge
   c. 3 - Average Knowledge
   d. 4 - Strong Knowledge
   e. 5 - Expert Knowledge
   f. Prefer not to answer
2. On a scale of 1 to 5, how much knowledge do you have of Psychosocial Concerns (i.e. depression, anxiety, fear of recurrence, finances, work, family needs, communication and relationships, etc.)?
   a. 1 - No Knowledge
   b. 2 - Slight Knowledge
   c. 3 - Average Knowledge
   d. 4 - Strong Knowledge
   e. 5 - Expert Knowledge
   f. Prefer not to answer
3. On a scale of 1 to 5, how much knowledge do you have of Nutrition-Based Concerns (i.e. weight management, vitamins and supplements, dietary restrictions, nutrition impact symptoms, etc.)?
   a. 1 - No Knowledge
   b. 2 - Slight Knowledge
   c. 3 - Average Knowledge
   d. 4 - Strong Knowledge
   e. 5 - Expert Knowledge
   f. Prefer not to answer
4. On a scale of 1 to 5, how much knowledge do you have of Rehabilitation and Exercise-Based Concerns (i.e. physical activity, physical limitations, fatigue, osteopenia/osteoporosis, lymphedema, peripheral neuropathy, etc.)?
   a. 1 - No Knowledge
   b. 2 - Slight Knowledge
   c. 3 - Average Knowledge
   d. 4 - Strong Knowledge
   e. 5 - Expert Knowledge
   f. Prefer not to answer
5. On a scale of 1 to 5, how much knowledge do you have of Late Effects/Physical Symptom Management (i.e. fatigue, lymphedema, peripheral neuropathy, cardiovascular health, pulmonary health, cognitive impairment, sexual dysfunction, etc.)?
   a. 1 - No Knowledge
   b. 2 - Slight Knowledge
   c. 3 - Average Knowledge
   d. 4 - Strong Knowledge
   e. 5 - Expert Knowledge
   f. Prefer not to answer
6. On a scale of 1 to 5, how much knowledge do you have of Discussion about the cancer diagnosis (i.e. staging, histologic grade, lymph node status, risk of recurrence and metastasis, tumor markers, etc.)?
   a. 1 - No Knowledge
   b. 2 - Slight Knowledge
   c. 3 - Average Knowledge
   d. 4 - Strong Knowledge
   e. 5 - Expert Knowledge
   f. Prefer not to answer
7. On a scale of 1 to 5, how much knowledge do you have of discussing the treatment a patient received (i.e. chemotherapy, radiation therapy, surgery, immunotherapy, targeted therapy, etc.)?
   a. 1 - No Knowledge
   b. 2 - Slight Knowledge
   c. 3 - Average Knowledge
   d. 4 - Strong Knowledge
   e. 5 - Expert Knowledge
   f. Prefer not to answer
8. On a scale of 1 to 5, how much knowledge do you have of discussing long-term management and follow-up with a patient (i.e. use of national guidelines to direct appropriate follow up testing, cancer prevention and surveillance, healthy lifestyle promotion, etc.)?
   a. 1 - No Knowledge
   b. 2 - Slight Knowledge
   c. 3 - Average Knowledge
   d. 4 - Strong Knowledge
   e. 5 - Expert Knowledge
   f. Prefer not to answer

**Thank you for your participation.**
